# Referral patterns in the CKD.QLD Registry: a call for revisiting the definition of late referral

**DOI:** 10.1101/2025.11.18.25340530

**Authors:** Clyson Mutatiri, Angela Ratsch, Matthew McGrail, Sree Krishna Venuthurupalli, Srinivas Kondalsamy-Chennakesavan

## Abstract

**Background:** The rising burden of chronic kidney disease (CKD) requires reinforcing of the current mechanisms for early detection and intervention. Although ‘timely’ referral to a specialist nephrology service plays a crucial role for many patients in ensuring improved outcomes, the lack of widely accepted definitions of timely (and by implication, early and late) referral may limit such outcomes.

**Study aim:** To evaluate referral patterns among CKD.QLD Registry participants, focusing on the timing and appropriateness of referrals and their association with clinical outcomes.

**Methods:** We conducted a retrospective cohort study of adults (≥18 years) enrolled in the CKD.QLD Registry from seven public nephrology clinics between May 2011 and June 2018. Participants were followed until kidney replacement therapy (KRT), death, or study closure. All CKD stages were included to assess demographic and clinical characteristics at referral and associated outcomes. Late referral was defined as initiation of KRT within 12 months of the first nephrology visit among those who progressed to KRT.

**Results:** Among 3,775 participants (median age 65 years; 54.8% male; 8.7% Indigenous), hypertension (77.5%) and type 2 diabetes (44.9%) were the most common comorbidities. ESKD developed in 775 (20.5%) participants; 513 (66.2%) received KRT. Overall, 722 (19.1%) died, including 124 (17.2%) after starting KRT. Late referral occurred in 60 (11.7%) KRT participants, with 15 (25%) deaths, 3 within 12 months of KRT initiation. Most KRT patients (77.4%) and deaths (65.5%) were in the high-comorbidity group; among KRT-related deaths, 101 (81.5%) were high-comorbidity, and 351 (88.4%) were referred early.

**Conclusion:** Late referral rates were lower than previously reported. Comorbidity burden, rather than referral timing, was the stronger predictor of mortality. These findings highlight the need for a standardized definition of late referral that incorporates risk stratification.

## Introduction

Chronic kidney disease (CKD) is a globally recognised contributor to increasing mortality and morbidity, with an estimated worldwide prevalence of 13.4% [1]. The burden of CKD is disproportionately borne by low-income countries and socio-economically disadvantaged populations [2]. With increasing life expectancy, coupled with increasing prevalence of key risk factors such as diabetes mellitus (DM) and hypertension, CKD has emerged as a critical public health concern and drain on resources for health systems globally [3]. Early identification and intervention are crucial for slowing the progression of CKD and improving patient outcomes[4].

Early-stage CKD is predominately managed within primary care settings [5]. However, a subset of these patients will ultimately require referral to nephrology clinics for specialist input, particularly as their disease advances or if they develop complications [6]. Traditionally, the referral of individuals with CKD from primary care to specialist nephrology services has been guided by established protocols, such as those outlined by international bodies including Kidney Disease: Improving Global Outcomes (KDIGO) [7], Kidney Health Australia (KHA) [8], and other globally recognised guidelines. The timing of referral has historically been considered a critical factor influencing outcomes, with the evidence demonstrating that earlier referral allows for better preparation for kidney replacement therapy (KRT), optimised management of comorbidities, and improved survival [9]. Late referral generally refers to patients who had commenced KRT such as dialysis or kidney transplantation within a short period (ranging from 1 to 12 months) following their initial nephrology clinic visit [10]. While there is no universally agreed-upon timeframe, this operational time-based definition is widely utilised in clinical studies [11].

Multiple studies have reported that late referral is associated with worse clinical outcomes, including higher mortality rates, increased rates of hospitalisation, and poorer quality of life [12, 13]. These adverse outcomes have been attributed to insufficient pre-dialysis education, suboptimal management of anaemia, mineral bone disorder, and delayed preparation for vascular access or transplantation [9]. However, the landscape of CKD management has evolved considerably in recent years. Advances in the understanding of CKD pathophysiology, the introduction of a working system based on estimated glomerular filtration rate (eGFR) and albuminuria, and the recognition of albuminuria as an important marker for early detection and risk stratification have all contributed to improved care [14, 15]. The emergence of renoprotective pharmacological therapies, such as renin-angiotensin-aldosterone system (RAAS) inhibitors, sodium-glucose co-transporter-2 (SGLT2) inhibitors, and statins, has enabled primary care clinicians to modify the risk of CKD progression, cardiovascular disease (CVD), and mortality at an earlier stage [16]. As a result, risk factor modification and the application of guideline-based therapy are increasingly being implemented in primary care settings before referral to nephrology [4]. The focus has shifted from simply managing eGFR thresholds to a more holistic approach that includes blood pressure control, optimising glycaemic management in diabetes, and the use of agents that offer renoprotection [17, 18].

Given this evolution in CKD care, the question arises: does the timing of referral to nephrology clinics still matter as a determinant of outcomes, particularly death following the initiation of KRT? To address this, we conducted a retrospective analysis of participants enrolled in the Chronic Kidney Disease Queensland Registry (CKD.QLD) to evaluate whether late referral continues to impact outcomes in patients who progressed to KRT. Additionally, the study seeks to critically evaluate and potentially refine the current operational definition of late referral from primary care to nephrology services.

## Methods

### Data source and methods of data collection

The Chronic Kidney Disease in Queensland (CKD.QLD) was established as a collaborative of most public sector kidney care practices in Queensland in May 2011. The CKD.QLD data collection methods have been described by Venuthurapalli *et al*, briefly however, the main objective of the registry was to profile participants with CKD, promoting CKD surveillance, practice improvement and research within the kidney care practice network in the public health system in QLD [19]. Data linkage framework was used to centralise data captured by multiple mechanisms to an individual participant via a unique identifier. This enabled further characterisation of participants with CKD, providing a comprehensive view of the health services utilisation and costs and outcomes of people with CKD.

Adult participants aged 18 years and above with the diagnosis of CKD presenting to public ambulatory nephrology clinics in the QLD health (QH) system were consented to have their demographic and clinical data collected. More than 7,600 participants consented to be part of the Registry between June 2011 and June 2018. Excluded were patients on kidney replacement therapy (KRT) and those with acute kidney injury (AKI) unless they subsequently developed and met the diagnostic criteria for CKD. Participants were followed until the start of KRT, death or 30 June 2018. For the current study, we excluded participants whose date of referral to the nephrology clinic couldn’t be established.

The demographic variables recorded included age, gender, ethnicity and geographical location, identified by postcode, as well as laboratory and clinical variables and outcome measures. Variables of interest which were not already in the database were manually collected from integrated electronic Medical Record (ieMR), the Hospital Based Corporate Information System (HBCIS), other QLD health platforms, and from private pathology laboratories as appropriate, following granting of a Public Health Act (PHA) approval. Where only urine protein-to-creatinine ratio (PCR) and dipstick protein were available, the equations developed by Sumida et al were employed to calculate the predicted urine albumin-to-creatinine ratio (UACR)[20]. The eGFR was calculated from serum creatinine using the Chronic Kidney Disease Epidemiology Collaboration (CKD-EPI) equation). Data extraction was undertaken between 18 January 2021 and 15 June 2025 in compliance with ethics and PHA approvals.

### Study design and population

This retrospective cohort study utilised data from participants aged ≥18 years enrolled in the CKD.QLD Registry between 1 May 2011 and 30 June 2018 from seven public healthcare nephrology clinics. A description of participants and basic study outcomes were previously detailed in a published protocol [21].

### Outcomes and variables

Primary outcomes included progression to end-stage kidney disease (ESKD), commencement of KRT, and mortality before and after commencement of KRT. Demographic and clinical variables were investigated for their potential role as predictors or covariates of these outcomes. Referral timing to nephrology services was assessed and defined by the conventional method as the interval between the first encounter with the nephrology service and commencement of KRT. Late referral was defined as commencing KRT within 12 months of the first visit to the nephrology clinic, amongst those who progressed to KRT.

The kidney failure risk equation (KFRE) was applied to participants with CKD stages G3 to G5 at the time of referral and available urine albumin-to-creatinine ratio (UACR) data to calculate the risk of progression to KRT. Socio-Economic Indexes for Areas (SEIFA) Relative Socio-economic Disadvantage (IRSD) scores as produced by the Australian Bureau of Statistics were used to measure socioeconomic status of the enrolled participants by their residential postcode [22]. The IRSD deciles were regrouped into quintiles (lowest, low, middle, high, and highest) with the highest quintile representing the least disadvantaged postal areas and the lowest quintile representing the most disadvantaged postal areas [23]. Participants were further categorised into 3 levels of socio-economic status by combining the two lowest quintiles into the low socio-economic status group, the two highest quintiles into the high socio-economic status group, and the middle quintile into the middle socio-economic status group. Participants with a recorded BMI were divided into obese (BMI ≥30) and non-obese (BMI <30). Comorbidity burden was dichotomised into low (0 to 2 comorbidities) and high (3 or more comorbidities).

### Data Analysis

Descriptive statistics and basic inferential statistics were employed to summarise demographic and clinical characteristics, exploring basic data patterns and describing referral patterns. Categorical variables were presented as proportions (numbers and percentages) and continuous variables as means with standard deviations. Associations between time of referral and the outcomes of death and KRT initiation were explored using a Cox proportional hazards regression model. The initial multivariable model assessed the rate of mortality against the following predictors: age at referral, eGFR at referral, gender, indigenous status, socioeconomic status, primary renal disease (PRD), comorbidity score, and the comorbidities of DM, CVD, hypertension, coronary artery disease (CAD), and the timing of referral presented as late referral. The final model excluded variables with little univariate significance, (p-value of >0.2) and summarized hazard ratios (HR) and 95% confidence intervals (95% CIs). The Log-rank test was used to compare the difference of survival by referral groups, and the Kaplan-Meier survival curve was used to depict the survival differences between the late and early referred groups after KRT initiation. All analyses used STATA version 18 and *p <* 0.05 as the threshold for statistical significance.

### Ethical considerations

The CKD.QLD registry and the hospital record(s) were examined by the research team retrospective to the participants’ details being entered into CKD.QLD and their hospital admissions. Both CKD.QLD data and hospital records contained identifiable information essential for data linkage. At enrolment, participants provided informed consent for CKD.QLD, which included permission to access and link relevant clinical information, such as medical history, pathology reports, and prior hospital admissions. Due to the large number of participants, obtaining individual consent for this study was not feasible. Instead, researchers obtained approval from the CKD.QLD Data Custodian to use existing consent arrangements under HREC/15/QRBW/294 (ERM: 13429) (AM05). Ethics approval was granted by the Royal Brisbane and Women’s Hospital Human Research Ethics Committee in January 2021 (LNR/2020/QRBW/69707, 14/01/2021) to access hospital records using unit record numbers provided by CKD.QLD for matching, with the intention to publish the findings. The requirement for individual consent was waived by the Ethics Committee, as all participants had previously consented to the use of their data for research upon registry enrolment.

Additionally, a Public Health Act approval (QCOS/029817/ RD006802) was granted in August 2023 for the release of missing patient data held by the contributing Queensland Health Hospital and Health Services.

## Results

### Study sample characteristics

Figure 1 outlines the participant selection process. A total of 3,775 participants were included in the final analysis with median age of 65 years (mean 62.5). Of these, 2,068 (54.8%) were male and 327 (8.7%) identified as Indigenous Australians (Table 1).

**Table 1.** Baseline characteristics at the time of referral and outcomes.

| Characteristics | Total sample | Stage 1 | Stage 2 | Stage 3A | Stage 3B | Stage 4 | Stage 5 |
| --- | --- | --- | --- | --- | --- | --- | --- |
| Participants, n (%) | 3775 | 7.9% | 11.0% | 18.6% | 31.0% | 26.2% | 5.3% |
| **Demographics** |  |  |  |  |  |  |  |
| Age, mean | 62.5 | 48.2 | 57.6 | 62.5 | 65.3 | 65.1 | 64.3 |
| Gender, male n (%) | 2,069 (54.8) | 46.1 | 56.0 | 57.8 | 52.6 | 56.5 | 58.8 |
| Indigenous, n (%) | 327 | 32 | 49 | 52 | 86 | 89 | 19 |
| **SES Status** |  |  |  |  |  |  |  |
| Low status n (%) | 1806 (47.8) | 48.2 | 52.2 | 49.9 | 47.4 | 45.4 | 45.7 |
| Middle status n (%) | 1006 (26.7) | 25.9 | 24.5 | 25.6 | 26.8 | 28.1 | 28.1 |
| High status n (%) | 963 (25.5) | 25.9 | 23.3 | 24.5 | 25.8 | 26.5 | 26.1 |
| BMI, n (mean) | 2500 (31.7) | 31.6 | 32.2 | 31.5 | 31.9 | 31.6 | 30.9 |
| **Primary Renal disease** |  |  |  |  |  |  |  |
| DN, n (%) | 1,066 (28.2) | 27.6 | 30.8 | 29.9 | 26.9 | 27.8 | 28.6 |
| GN, n (%) | 473 (12.5) | 12.5 | 12.0 | 11.4 | 13.2 | 12.5 | 14.1 |
| GRD, n (%) | 183 (4.9) | 3.7 | 12.0 | 4.3 | 5.4 | 5.2 | 4.5 |
| RV, n (%) | 985 (26.1) | 23.9 | 28.4 | 28.6 | 27.0 | 23.1 | 25.6 |
| UNC, n (%) | 403 (10.7) | 10.8 | 8.4 | 9.8 | 11.4 | 12.2 | 6.5 |
| Other, n (%) | 665 (17.6) | 21.6 | 15.9 | 16.1 | 16.3 | 19.3 | 20.6 |
| **Comorbidities** |  |  |  |  |  |  |  |
| DM, n (%) | 1,694 (44.9) | 32.0 | 42.8 | 46.0 | 47.1 | 46.3 | 44.2 |
| HTN, n (%) | 2,926 (77.5) | 77.7 | 76.7 | 74.8 | 77.4 | 80.7 | 78.9 |
| CVD, n (%) | 1,538 (40.7) | 21.2 | 32.2 | 41.1 | 43.4 | 47.4 | 37.7 |
| CAD, n (%) | 886 (23.5) | 16.8 | 19.2 | 23.3 | 25.0 | 26.1 | 23.5 |
| Obesity, n (%) | 1,366 (54.6) | 51.2 | 56.6 | 53.5 | 56.9 | 54.0 | 49.1 |
| **Comorbidity Score** |  |  |  |  |  |  |  |
| Low comorbidity, n (%) | 1223 (32.4) | 34.0 | 33.2 | 37.1 | 29.8 | 32.0 | 32.4 |
| High comorbidity, n (%) | 2552 (67.6) | 66.0 | 66.8 | 62.9 | 70.2 | 68.0 | 67.6 |
| **Laboratory Parameters** |  |  |  |  |  |  |  |
| eGFR, mean | 42.4 | 90 | 71.5 | 51.0 | 36.7 | 22.9 | 11.4 |
| UACR, mean | 56.1 | 44.2 | 68.5 | 48.6 | 44.5 | 66.5 | 86.8 |
| **Outcomes** |  |  |  |  |  |  |  |
| ESKD, n (%) | 775 (20.5) | 5.7 | 11.8 | 12.4 | 16.4 | 34.4 | 54.8 |
| KRT, n (%) | 513 (13.6) | 3.7 | 8.4 | 8.8 | 10.6 | 23.4 | 34.7 |
| Total death, n (%) | 722 (19.1) | 5.7 | 10.6 | 15.1 | 18.2 | 27.1 | 36.7 |
| Death without KRT, n (%) | 598 (15.8) | 5.7 | 10.1 | 12.7 | 15.7 | 21.1 | 29.2 |

SD: standard deviation; BMI: body mass index; DN: diabetic nephropathy; GN: glomerulonephritis; GRD: genetic renal disease; RV: renovascular disease; UNC: unknown cause; DM: diabetes mellitus; HTN: hypertension; CVD: cardiovascular disease; CAD: coronary artery disease; eGFR: estimated glomerular filtration rate; UACR: urine albumin-to-creatinine ratio; ESKD: end-stage kidney disease; KRT: kidney replacement therapy

Socioeconomic status, as measured by the IRSD, revealed that a significant portion of the participants (1,806, 47.8%) lived in the most disadvantaged postal areas (low socioeconomic status) (Table 1). This group represented the largest proportion of participants across all stages of CKD (Table 1).

At the time of referral, nearly half of the participants, 1,872 (49.6%) were in stage G3 CKD (G3A and G3B), whereas 713 (18.9%) were in stages 1 or 2 (eGFR of ≥60ml/min/1.73m^2^). CKD stages G4 and 5 accounted for 1,190 (31.5%) participants, predominately G4 (Table 1). UACR data were available for 2,605 (69%) participants, with 920 (35.3%) exhibiting a UACR of ≥ 30mg/mmol (Table S1).

There were 17 types of comorbidities recorded, with the number of comorbidities ranging from 0 to 15 per participant. A high proportion met the high comorbidity score criteria (range 63%-70%) in all stages of CKD (Table 1). There were twice as many participants in the high comorbidity group (67.6%) of whom 1,737 (68.1%) had an eGFR of ≥30 ml/min at the time of referral (Table S2). Hypertension was the most common comorbidity, present in 77.5% of participants, followed by DM and cardiovascular disease (CVD), 44.9% and 40.7% respectively (Table 1).

### Referral thresholds and outcomes

Around one third of participants (1,194, 31.6%) fulfilled the KHA eGFR threshold (<30ml/min) for specialist referral (Table S2). Among the 2,581 (68.4%) who did not fulfill the eGFR threshold, 216 (8.4%) progressed to KRT while 331 (12.8%) died without KRT (Table S2). Similarly, though 1,685 (64.7%) participants did not fulfil the UACR threshold of ≥30 mg/mmol for referral, of these 191 (11.3%) progressed to KRT while 221 (13.1%) died without KRT (Table S1).

Of the 2,605 participants with both eGFR and UACR data, 371 (14.2%) met both referral criteria, of which 150 (40.4%) subsequently requiring KRT (Table S3). Conversely, 1,203 (46.2%) did not meet either threshold for referral, yet 79 (6.6%) of these went on to require KRT at some point during the period of follow up (Table S3).

### Mortality and progression to ESKD

During the follow up period, 722 (19.1%) participants died, with the majority (598, 82.8%) dying prior to commencing KRT, while 775 (21%) participants progressed to ESKD, of whom 513 (66.2%) received KRT (Table 1). Nearly half of all deaths (356, 49.3%) occurred among participants from the low socioeconomic status group (Figure 2).

The Cox proportional hazards regression model indicated that participants who had been under nephrology care for less than 12 months before starting KRT (late referral) had a 54% lower hazard rate of mortality compared to those who had received care for more than 12 months (HR <12 months, 0.46; 95% CI, 0.27 to 0.79; p = 0.01) (Table 2). The Log-rank test showed a statistically significant difference in survival between the referral groups (p = 0.002) (Table S4). The Kaplan-Meier survival curve shows longer survival after KRT initiation in the late referred group (Figure 3). Advancing age was associated with increased mortality rate, with each 5-year increment corresponding to a 15% increase in the hazard of death (HR□=□1.15). Additionally, the presence of cardiovascular disease was associated with a nearly twofold increase in mortality rate (HR□=□1.90) (Table 2).

**Table 2.** Final Cox regression model.

| Predictor | Hazard ratio | [95% CI] | p-value |
| --- | --- | --- | --- |
| Age at referral | 1.15 | 1.07 – 1.24 | <0.01 |
| Cardiovascular Disease | 1.9 | 1.31 – 2.75 | <0.01 |
| Late referral | 0.46 | 0.27 – 0.79 | 0.01 |

CI: confidence interval

### Referral timing and comorbidity

Of the 513 participants who commenced KRT, 60 (11.7%) were classified as late referrals. Among these, 32 (53.3%) were from low socioeconomic group, compared to 20 (33.3%) from high socioeconomic group (Table 3).

**Table 3.**
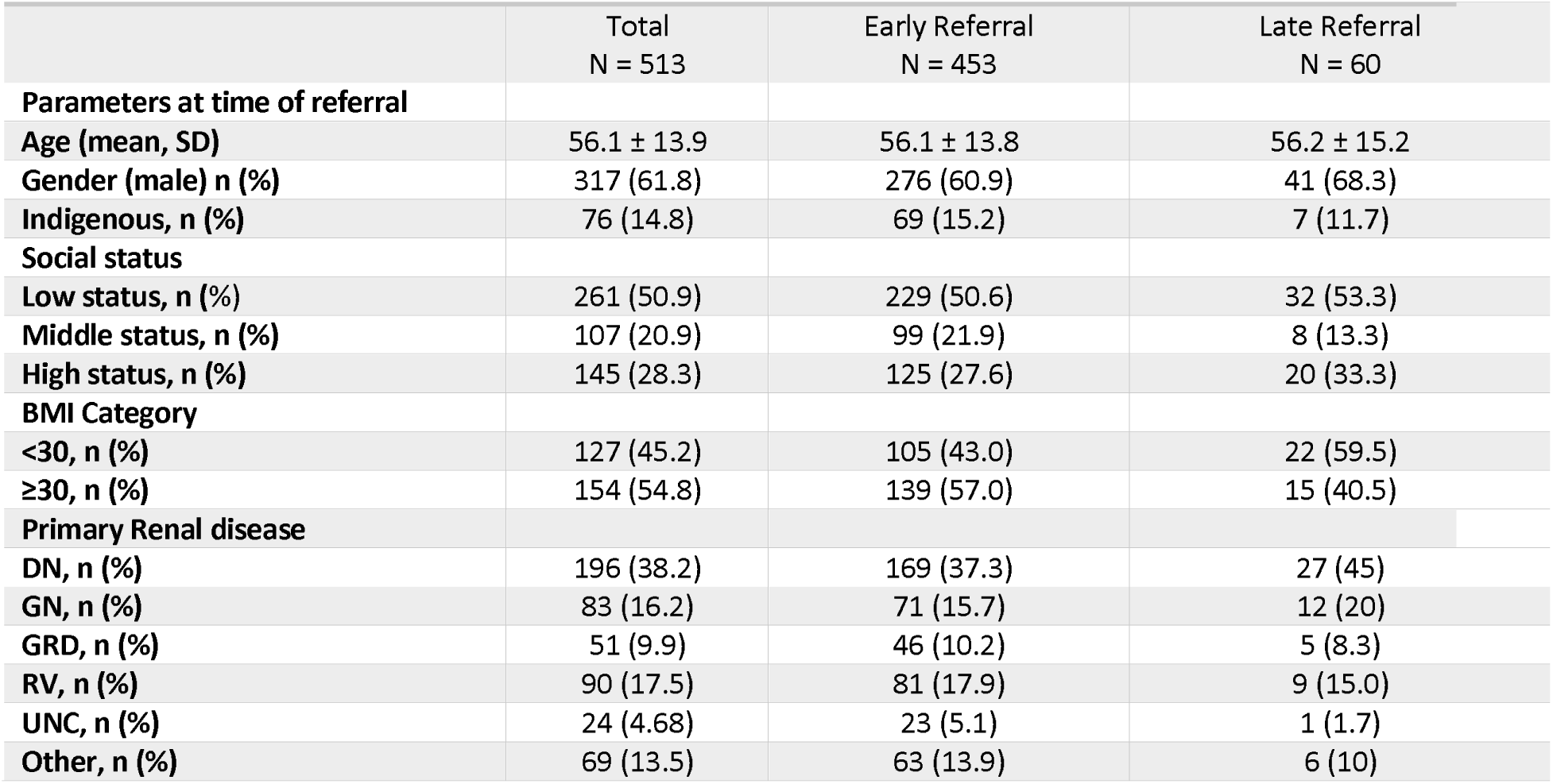
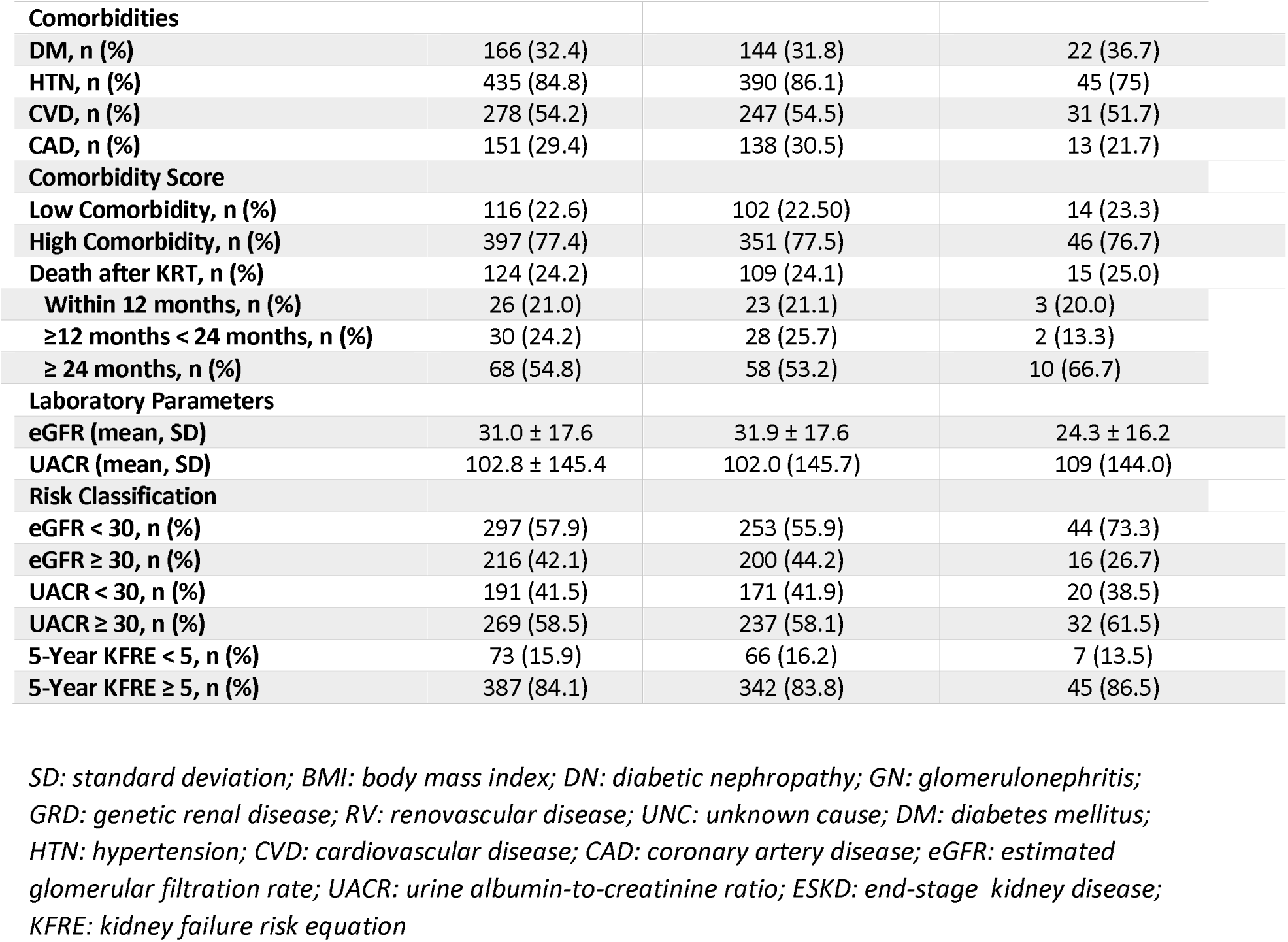
Characteristics of participants who progressed to KRT.

|  | Total<br>N = 513 | Early Referral<br>N = 453 | Late Referral<br>N = 60 |
| --- | --- | --- | --- |
| **Parameters at time of referral** |  |  |  |
| Age (mean, SD) | 56.1 ± 13.9 | 56.1 ± 13.8 | 56.2 ± 15.2 |
| Gender (male) n (%) | 317 (61.8) | 276 (60.9) | 41 (68.3) |
| Indigenous, n (%) | 76 (14.8) | 69 (15.2) | 7 (11.7) |
| **Social status** |  |  |  |
| Low status, n (%) | 261 (50.9) | 229 (50.6) | 32 (53.3) |
| Middle status, n (%) | 107 (20.9) | 99 (21.9) | 8 (13.3) |
| High status, n (%) | 145 (28.3) | 125 (27.6) | 20 (33.3) |
| **BMI Category** |  |  |  |
| <30, n (%) | 127 (45.2) | 105 (43.0) | 22 (59.5) |
| ≥30, n (%) | 154 (54.8) | 139 (57.0) | 15 (40.5) |
| **Primary Renal disease** |  |  |  |
| DN, n (%) | 196 (38.2) | 169 (37.3) | 27 (45) |
| GN, n (%) | 83 (16.2) | 71 (15.7) | 12 (20) |
| GRD, n (%) | 51 (9.9) | 46 (10.2) | 5 (8.3) |
| RV, n (%) | 90 (17.5) | 81 (17.9) | 9 (15.0) |
| UNC, n (%) | 24 (4.68) | 23 (5.1) | 1 (1.7) |
| Other, n (%) | 69 (13.5) | 63 (13.9) | 6 (10) |
| **Comorbidities** |  |  |  |
| DM, n (%) | 166 (32.4) | 144 (31.8) | 22 (36.7) |
| HTN, n (%) | 435 (84.8) | 390 (86.1) | 45 (75) |
| CVD, n (%) | 278 (54.2) | 247 (54.5) | 31 (51.7) |
| CAD, n (%) | 151 (29.4) | 138 (30.5) | 13 (21.7) |
| **Comorbidity Score** |  |  |  |
| Low Comorbidity, n (%) | 116 (22.6) | 102 (22.50) | 14 (23.3) |
| High Comorbidity, n (%) | 397 (77.4) | 351 (77.5) | 46 (76.7) |
| Death after KRT, n (%) | 124 (24.2) | 109 (24.1) | 15 (25.0) |
| Within 12 months, n (%) | 26 (21.0) | 23 (21.1) | 3 (20.0) |
| ≥12 months < 24 months, n (%) | 30 (24.2) | 28 (25.7) | 2 (13.3) |
| ≥ 24 months, n (%) | 68 (54.8) | 58 (53.2) | 10 (66.7) |
| **Laboratory Parameters** |  |  |  |
| eGFR (mean, SD) | 31.0 ± 17.6 | 31.9 ± 17.6 | 24.3 ± 16.2 |
| UACR (mean, SD) | 102.8 ± 145.4 | 102.0 (145.7) | 109 (144.0) |
| **Risk Classification** |  |  |  |
| eGFR < 30, n (%) | 297 (57.9) | 253 (55.9) | 44 (73.3) |
| eGFR ≥ 30, n (%) | 216 (42.1) | 200 (44.2) | 16 (26.7) |
| UACR < 30, n (%) | 191 (41.5) | 171 (41.9) | 20 (38.5) |
| UACR ≥ 30, n (%) | 269 (58.5) | 237 (58.1) | 32 (61.5) |
| 5-Year KFRE < 5, n (%) | 73 (15.9) | 66 (16.2) | 7 (13.5) |
| 5-Year KFRE ≥ 5, n (%) | 387 (84.1) | 342 (83.8) | 45 (86.5) |

SD: standard deviation; BMI: body mass index; DN: diabetic nephropathy; GN: glomerulonephritis; GRD: genetic renal disease; RV: renovascular disease; UNC: unknown cause; DM: diabetes mellitus; HTN: hypertension; CVD: cardiovascular disease; CAD: coronary artery disease; eGFR: estimated glomerular filtration rate; UACR: urine albumin-to-creatinine ratio; ESKD: end-stage kidney disease; KFRE: kidney failure risk equation

Most participants who started KRT (397, 77.4%) and most who died (473, 65.5%) were in the high comorbidity group (Table 2). Of those who died after starting KRT, (101, 81.5%) were from the high comorbidity group, most of whom (351, 88.4%) had been referred early (Table S5). Only 15 (12.1%) of those who died post-KRT were late referrals, with 3 (20%) of these dying in the first 12 months of starting KRT (Table 3). These findings suggest that comorbidity burden rather than late referral is a more robust predictor of death.

### CKD stage and risk of progression

Participants in CKD stages 1 to 3 were found to be at greater risk of death before KRT than progress to KRT (55.4% vs 43.5%), whereas those in stages 4 & 5 were more likely to initiate KRT than die before KRT (56.5% vs 44.6%) (Table 1). Overall, mortality rates increased with advancing CKD stages (Figure S1)

### KFRE application and predictive accuracy

Among the 1,743 participants who did not fulfil the KHA eGFR criteria for referral and whose UACR was available, application of the 5-year KFRE at 3% & 5% thresholds identified 708 (40.6%) and 509 (29.2%) as high risk, respectively. Of these, 144 (20.3%) and 126 (24.8%) progressed to KRT, while 91 (12.9%) and 65 (12.8%) died without KRT (Table S6).

When applied to participants who met the KHA eGFR referral threshold, the KFRE at both 3% and 5% thresholds, demonstrated similar predictive accuracy for KRT progression (99.6% and 96.6% respectively). The observed risks for KRT and death were also similar (KRT:31.0% and 31.9%; death:15.6% and 14.9% (Table S7).

These findings suggest that the eGFR threshold alone is as effective as the 5-year KFRE in identifying individuals at risk of progressing to KRT as well as those who died without KRT. However, the KFRE enabled identification of additional high-risk participants who would otherwise be missed using the eGFR criteria alone.

## Discussion

In our study, we found that participants who were in the care of nephrology clinics for more than 12 months before commencing KRT (i.e. usually considered ‘timely’ referral) exhibited lower survival rates than those who were referred within 12 months of KRT initiation (i.e., ‘late’ referral). This finding contrasts with prior studies that have reported poorer survival rates in those patients referred late using the same 12-month threshold. Previous studies have found that late referral is associated with increased mortality and poorer outcomes after starting KRT [10, 11]. The presumed benefits of early referral included better preparation for dialysis or transplantation, timely management of complications, and improved patient education. However, these studies often predate the widespread adoption of contemporary CKD management strategies [12, 24].

These results suggest that the duration of nephrology care alone may not be a sufficient explanatory factor for survival after initiation of KRT. Previous research has highlighted that even patients who have been under nephrology care for longer periods may still undergo urgent start dialysis [25–27]. Moreover, nephrology care does not always ensure optimal dialysis preparation. For example, Hamadahet al. [28] found that only 30.8% of patients under nephrology care for more than 12 months started haemodialysis with an arteriovenous fistula (AVF). Similarly, Mendelssohnet al. [29] demonstrated that many patients who were followed by a nephrologist experienced suboptimal dialysis commencement, defined as using a temporary central dialysis catheter access for haemodialysis or peritoneal dialysis immediately after placement of a PD catheter. A follow-up study by the same group replicated the results of their study by demonstrating a 56.4% rate of suboptimal starts in early referred patients [30].

Commencing dialysis with a temporary catheter has often been cited as a key reason why patients fare poorly after starting dialysis [31]. In our analysis, we chose the 12-months cut-off to define late referral to eliminate vascular access as a potential cause of poor outcomes after starting dialysis, arguing that 12 months would be long enough for the nephrology clinics to facilitate the creation of permanent vascular access before the start of dialysis. While it can be argued that timing of referral alone (i.e., duration of pre-dialysis care) cannot be sufficient to account for the superior outcomes observed in early referred patients, it is also instructive to acknowledge that a longer duration of pre-dialysis care creates opportunities for an increased number of visits to the nephrology clinics and allows for more time for effective treatment and correction of anaemia and other complications of CKD, hence likely lowering the risk of death. Moreover, frequent visits to the nephrology clinic and early exposure to multidisciplinary care clinics have been demonstrated to lead to timely dialysis initiation. Early exposure allows adequate and comprehensive predialysis education which in turn promotes supported modality selection and timely permanent access creation [32, 33].

We observed a high burden of comorbidities in all participants, most of whom had an eGFR of ≥30 ml/min at the time of referral. This trend was maintained in participants who progressed to KRT; a higher burden of comorbidities was observed in those who were referred early (commenced KRT after 12 months of referral) and in those who died after commencing KRT. These results suggest that participants who had more comorbid conditions were more readily referred early to nephrology practice rather than stable patients, whose main reason of referral would have been uncomplicated CKD. Preexisting comorbidities have been reported to predict higher mortality rates in patients starting dialysis [34]. In a prospective cohort study of patients with CKD stage G5 and delayed initiation of haemodialysis, patients with a high and very high Charlson Comorbidity Index (CCI) were found to have an increased risk for all-cause mortality, with mortality risk increasing progressively as the CCI score increased [35]. High comorbidity rate therefore appears to have been the major determinant of mortality after KRT initiation, superseding the impact of late referral to nephrology services, which in our study did not predict mortality.

Low socioeconomic status was a key characteristic observed in all stages of CKD and was also a predictor of both progression to ESKD and mortality before and after initiation of KRT. Studies have previously demonstrated low socioeconomic status to be a significant risk factor for CKD progression and mortality, leading to higher rates of CVD and kidney failure, and ultimately, increased all-cause mortality [36]. Factors that have been identified as drivers of CKD progression and death in individuals from low socioeconomic background include delayed diagnosis due to poor access to healthcare, unfavourable living conditions, poor nutrition, and higher burden of chronic diseases [37].

On assessing compliance with the KHA referral criteria, we observed that 46.3% of participants whose UACR was available did not fulfill either the eGFR or UACR thresholds for referral, which means that they would have been either referred for reasons other than the eGFR or UACR thresholds, or they may have been referred inappropriately and hence could have been managed in primary care. In a similar retrospective cohort study of CKD patients over 14 years, Ghimireet al. [38] observed guideline discordant rate of 59% and concluded that many of the referred patients being seen by nephrologists might have had mild CKD that could have been safely managed in primary care. Singhet al. [39] reported that adherence to the KDIGO guidelines would result in a supply-demand mismatch and concluded that implementing the guidelines may not be feasible. Kielet al. [40] came to a similar conclusion, reporting that implementing KDIGO criteria would lead to more than double increase in referral rate compared to actual referral. It has therefore been long argued that most current referral criteria are not designed to accurately distinguish between high and low risk patients. In our study, we observed that a significant proportion of participants who did not fulfill the KHA eGFR threshold for referral were deemed high risk when the 5-year KFRE was applied at both the 3% and 5% thresholds. Additionally, when applied to participants who did not fulfill the eGFR threshold for referral, the 5-year KFRE better identified participants who progressed to KRT than who died.

The period when the participants in our study were followed up in CKD.QLD Registry in relation to worldwide kidney care practice patterns is important. Substantial improvements in the management of CKD in primary care settings may explain our study’s differing results. Enhanced education of primary care providers, improved disease definition and staging, and the integration of guideline-based therapies have collectively contributed to better patient outcomes. The widespread use of new pharmacological agents, particularly those with proven renoprotective and cardioprotective effects, has further lessened the reliance on specialist referral for optimal care.

These results suggest that we might be seeing the impact of improved CKD care in terms of early detection and timely intervention within primary care to mitigate the risk of late referral. With the advent of therapies capable of delaying the need for dialysis by decades, the necessity for routine referral based solely on eGFR thresholds or arbitrary timing will need to be reevaluated. Referral to nephrology may be best reserved for high-risk patients, identified through validated risk prediction models like the KFRE, or those with complex needs such as glomerulonephritis, vasculitis, genetic kidney diseases, or advanced CKD (stages G4 and G5) requiring specialised interventions (e.g., phosphate binders, erythropoiesis-stimulating agents, or preparation for KRT). Most patients referred to nephrology clinics are already receiving optimised medical therapy, making the incremental benefit of specialist care less pronounced in the early stages of CKD.

One of the key strengths of our study is that it includes important baseline characteristics of participants at the time they were referred, unlike many existing studies on CKD referral patterns that focus only on patient data at the start of KRT. Additionally, we were able to calculate the UACR for a substantial number of participants, which allowed us to use the KFRE to compare predicted outcomes with actual results. However, the retrospective nature of the study meant that some clinical and laboratory data were missing from the Registry, limiting the depth of our analysis. Other limitations included being unable to determine the reasons for referral to nephrology clinics, making it difficult to assess whether participants who didn’t meet the eGFR and UACR criteria were referred appropriately. Furthermore, the Registry did not record the type of vascular access used at the start of KRT, preventing us from evaluating its potential effect on post-KRT mortality.

### The case for a new definition of late referral: A risk-based approach

We propose that risk stratification be incorporated into the definition of the timeliness of referral of individuals with CKD from primary care to specialist nephrology services, and that the definition be applicable at the point of initial nephrology consultation. This prospective approach will enable timely interventions to mitigate adverse outcomes. The current retrospective definition, based on variable intervals between the first nephrology visit and initiation of KRT, lacks universal agreement, with timeframes ranging from 1 to 12 months. This lack of a universally accepted time interval has compromised the integrity of systematic reviews which have been conducted on the topic due to the heterogeneity of the time frames used in the many retrospective cohort studies available in the literature. Moreover, retrospective definitions offer little guidance to primary care physicians seeking optimal referral timing, nor do they assist nephrologists in assessing referral appropriateness at the time of the first clinic visit. Having this knowledge upfront will enable clinicians to streamline their delivery of care and resources according to specific goals of care, that is, whether initiating KRT preparation or focusing on slowing CKD progression and reducing cardiovascular risk and death.

### Conclusions

Our analysis shows that the rate of late referral in our sample was lower than the rates that have been reported in previous studies, suggesting that in the current era of enhanced CKD management at the primary care level, the timing of referral to nephrology services is no longer a significant determinant of post-KRT mortality. Instead, high comorbidity burden and low social status seemed to be stronger predictors of mortality after KRT initiation. Other results found that the accuracy of the KHA eGFR threshold of <30ml/min was comparable to the KFRE 5-year risk of 3% and 5% in identifying participants who required KRT or died, however, it missed a significant proportion of high-risk participants who later progressed to KRT and death.

We conclude that the timing of referral may no longer be reliable in determining outcomes for patients with CKD, given the advancements in primary care management and the availability of effective renoprotective therapies and suggest that efforts be made to develop a more universally accepted definition of late referral of patients with CKD from primary care to specialist nephrology that incorporates risk stratification. We also encourage the empowering of the primary care with targeted education programs with emphasis on guideline adherence and incorporation of renoprotective agents at primary care level so that referral to secondary care can be reduced.

## Author Declaration

The authors declare no competing interests.

## Author contributions

**Conceptualization**: Clyson Mutatiri, Sree Krishna Venuthurupalli, Srinivas Kondalsamy-Chennakesavan, Angela Ratsch, and Matthew McGrail.

**Methodology**: Clyson Mutatiri, Sree Krishna Venuthurupalli, Angela Ratsch Srinivas Kondalsamy-Chennakesavan, Matthew McGrail.

**Writing - original draft preparation**: Clyson Mutatiri, Matthew McGrail, Srinivas Kondalsamy-Chennakesavan, Angela Ratsch, Sree Krishna Venuthurupalli.

**Writing - review and editing**: Clyson Mutatiri, Matthew McGrail, Angela Ratsch, Srinivas Kondalsamy-Chennakesavan, Sree Krishna Venuthurupalli.

## Data Availability

All relevant data are within the manuscript and its Supporting Information files.

## Acknowledgments

We would like to thank the CKD.QLD Registry for granting us the approval of access to CKD.QLD Registry Patient Data and support for the progression of this research. We would also like to acknowledge the “in-kind” support provided by Wide Bay Hospital and Health Service to the researchers and for relevant resources including computers, telephone, and office space for the duration of this project. This study was partly supported by the Australian Government Department of Health under the Rural Health Multidisciplinary Training Programme.

## Supporting information

**S1Table** UACR categories

**S2 Table** eGFR categories

**S3 Table** KRT by eGFR and UACR categories

**S4 Table** Log-rank test for equality of survivor functions by referral status

**S5 Table** Comorbidity score by KRT death and referral status

**S1 Fig** Distribution of death without KRT by CKD stage

**S6 Table** Comparison of eGFR threshold and KFRE Subgroup of participants with eGFR of ≥30ml/min

**S7 Table** Comparison of eGFR threshold and KFRE Subgroup of participants with eGFR of <30ml/min

---

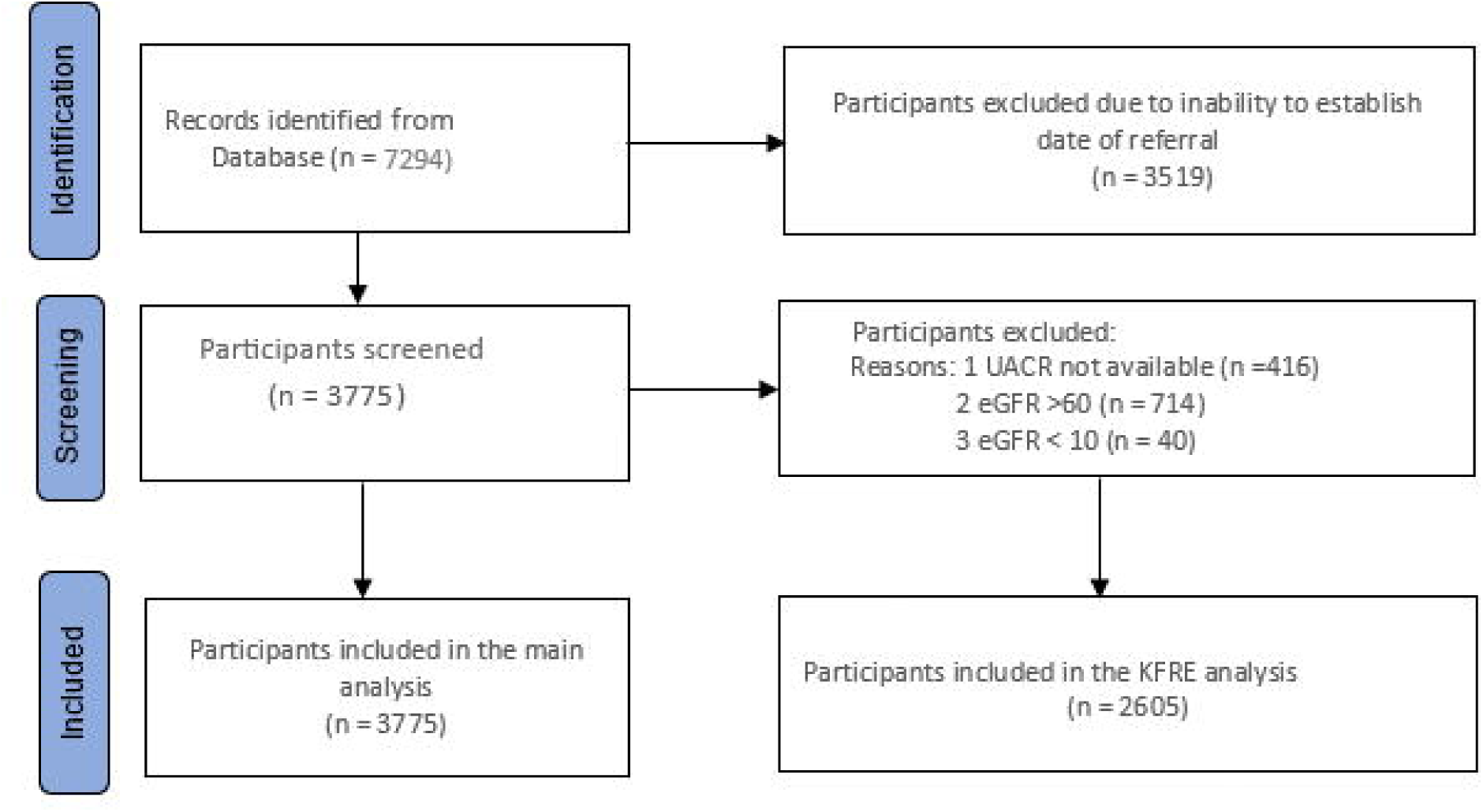

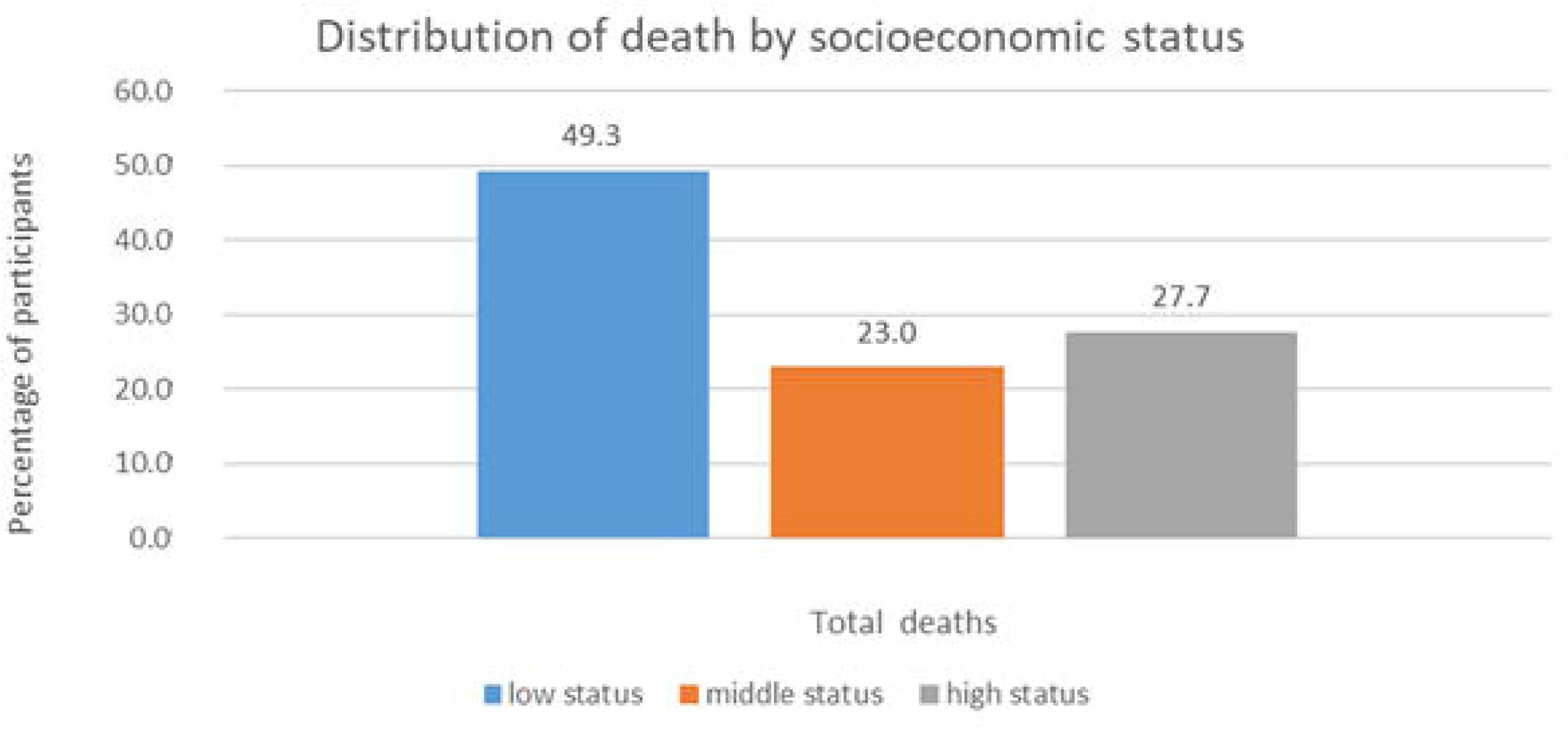

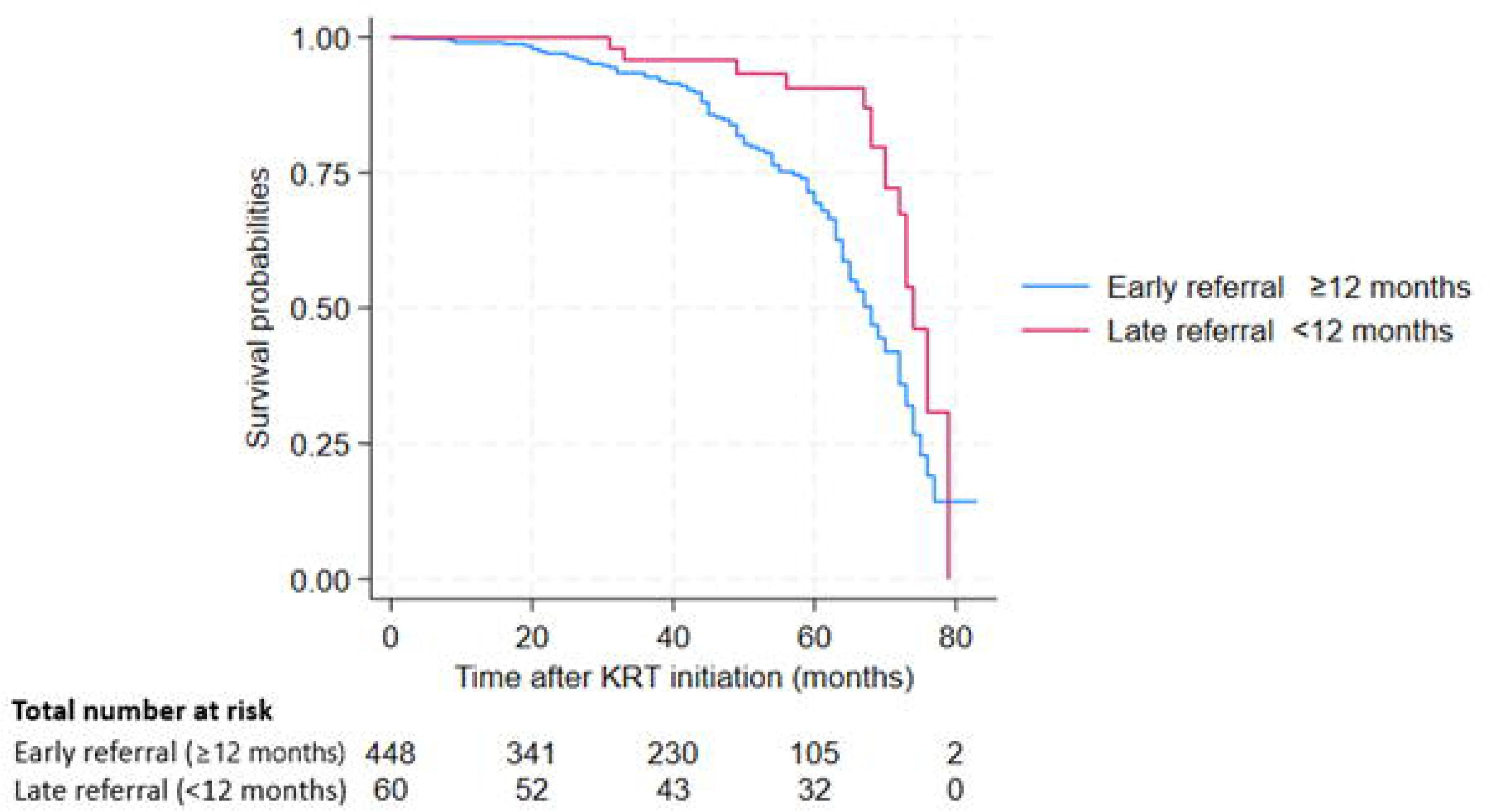

