## Supporting information for "Referral patterns in the CKD.QLD Registry: a call for revisiting the definition of late referral"

**S1Table** UACR categories

| KHA threshold | N = 2605 | KRT | Death without KRT |
| --- | --- | --- | --- |
| UACR <30 | 1685 (64.7%) | 191 (11.3%) | 221 (13.1%) |
| UACR ≥30 | 920 (35.3%) | 216 (8.4%) | 331 (12.8%) |

**S2 Table** eGFR categories

| KHA threshold | All participants | KRT | Death without KRT | Low comorbidity | High comorbidity |
| --- | --- | --- | --- | --- | --- |
| eGFR <30 | 1194 (31.6%) | 297 (24.9%) | 267 (22.4%) | 379 (31.0%)) | 815 (31.9%) |
| eGFR ≥30 | 2581 (68.4%) | 216 (8.4%) | 331 (12.8%) | 844 (69.0%) | 1737 (68.1%) |
| Total | 3775 | 513 | 598 | 1223 | 2552 |

**S3 Table** KRT by eGFR and UACR categories

|  | UACR category (number of KRT) | | |
| --- | --- | --- | --- |
| eGFR | <30 mg-mmol | >=30 mg-mol | Total |
| <30 ml-min | 482 (112) | 371 (150) | 853 (262) |
| >=30 ml-min | 1203 (79) | 549 (119) | 1752 (198) |
| Total | 1685 (191) | 920 (269) | 2605 (460) |

**S4 Table** Log-rank test for equality of survivor functions by referral status

| Timing of referral | Observed events | Expected events |
| --- | --- | --- |
| ≥12 months \| | 109 | 95 |
| <12 months | 15 | 29 |
| Total | 124 | 124 |

*chi2(1) =	9.21; Pr > chi2 = 0.0024*

**S5 Table** Comorbidity score by KRT death and referral status

| Comorbidity score | KRT Death | | Referral Status | |
| --- | --- | --- | --- | --- |
|  | No | Yes | ≥ 12 months | < 12 months |
| Low | 93 | 23 | 102 | 14 |
| High | 296 | 101 | 351 | 46 |
| Total | 389 | 124 | 453 | 60 |

**S6 Table** Comparison of eGFR threshold and KFRE Subgroup of participants with eGFR of ≥30ml/min

| N = 1743 | | KFRE <3% | KFRE ≥3% | KFRE <5% | KFRE ≥5% |
| --- | --- | --- | --- | --- | --- |
| Baseline | | 1035 (59.4%) | 708 (40.6%) | 1234 (70.8%) | 509 (29.2%) |
| KRT | Yes | 54 (5.2%) | 144 (20.3%) | 72 (5.8%) | 126 (24.8%) |
|  | No | 981 (94.8%) | 564 (79.7%) | 1162 (94.2%) | 383 (75.2%) |
| Death | Yes | 117 (11.3%) | 91 (12.9%) | 143 (11.6%) | 65 (12.8%) |
|  | No | 918 (88.7%) | 578 (81.6%) | 1077 (88.4%) | 444 (87.2%) |

**S7 Table** Comparison of eGFR threshold and KFRE Subgroup of participants with eGFR of <30ml/min

| N = 847 | | KFRE <3% | KFRE ≥3% | KFRE <5% | KFRE ≥5% |
| --- | --- | --- | --- | --- | --- |
| Baseline | | 16 (3%) | 844 (99.6%) | 29 (3.4%) | 818 (96.6%) |
| KRT | Yes | 0 (0%) | 262 (31.0%) | 1 (3.4%) | 261 (31.9%) |
|  | No | 16 (100%) | 582 (69.0%) | 28 (96.6%) | 557 (68.1%) |
| Death | Yes | 0 (0%) | 132 (15.6) | 10 (34.5%) | 122 (14.9%) |
|  | No | 3 (100%) | 712 (84.4%) | 19(65.5%) | 696 (85.1%) |
