## Supplementary figures and images for "Referral patterns in the CKD.QLD Registry: a call for revisiting the definition of late referral"

### S1Fig

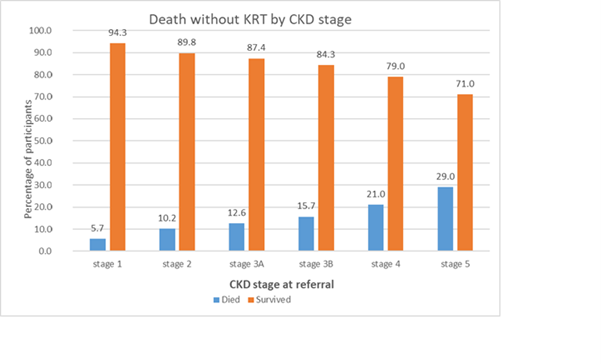
